# COVID-19 hospitalizations in the Netherlands, 2023-2024: disease burden and vaccine effectiveness

**DOI:** 10.64898/2026.02.12.26346177

**Authors:** Brechje de Gier, Bente Smagge, Annika van Roon, Irene Veldhuijzen, Pieter de Boer, Mirjam Knol, Susan Hahné, Hester de Melker

## Abstract

Since the cessation of real-time monitoring of COVID-19 hospitalizations in early 2024, the burden of and vaccine effectiveness (VE) against severe COVID-19 in the Netherlands was largely unknown. Recently, hospitalization data from 2024 were made available for the purpose of monitoring and evaluating the COVID-19 vaccination campaigns. These data were linked to the population registry, vaccination registry and healthcare use data (for classification into medical risk groups).

We analyzed the number and incidence of COVID-19 hospitalizations in 2023 and 2024 by age and medical risk group. VE against hospitalisation of the autumn booster of 2023 (by time since vaccination, 25 September 2023 to 16 September 2024) and of the autumn booster of 2024 (16 September to 31 December 2024) were estimated by medical risk group among persons aged 60 years and older using Cox proportional hazards models with calendar time as underlying time scale and vaccination status as time-varying exposure. Models were adjusted for age, sex, region and household socio-economic status.

From around age 60 onward, intermediate and high medical risk groups had a markedly higher incidence than younger age groups, increasing with age. Persons in the low medical risk group had a low incidence up to the age of 80. In 2024, incidence was lower than in 2023. For both autumn booster rounds, estimated VE against hospitalisation was moderate at 55-67% in the first 3 months post-vaccination. In the high medical risk group, 2023 VE decreased fast and was no longer significant at 6 months post-vaccination. For both years, estimates of the number of averted hospitalizations and number needed to vaccinate to prevent one hospitalization indicated that significant health benefit can be achieved by vaccinating the intermediate and high medical risk groups aged 60 years and older. Efforts to increase the moderate vaccine uptake among risk groups could potentially prevent a considerable disease- and healthcare burden.

**Highlights:**

- In 2023 and 2024, incidence of COVID-19 hospitalization was highest among medical risk groups aged 60 years and older, despite vaccination campaigns.
- Estimated VE against hospitalisation of the 2023 and 2024 autumn booster campaigns was moderate (55-67%) in the first year-quarter post-vaccination among persons aged 60 years and older.
- Estimated VE of the 2023 autumn booster decreased over the year, and faster among persons with a medical risk condition. Data availability precluded estimates of 2024 VE beyond the first 3 months since the start of the campaign.
- Despite lower and waning VE, the estimated number needed to vaccinate to prevent one COVID-19 hospitalization was much lower among intermediate and high medical risk groups compared with the low medical risk group.

## Introduction

After the pandemic phase of COVID-19, annual autumn booster vaccination campaigns to prevent severe COVID-19 were implemented in the Netherlands, targeting persons aged 60 years and older and specific risk groups. Monitoring of the burden of COVID-19 hospitalizations and vaccine effectiveness (VE) against COVID-19 hospitalization was based on near real-time data provided by the National Intensive Care Evaluation (NICE) registry in the period 2020-2023 [1, 2]. After the discontinuation of this comprehensive NICE COVID-19 registration, data to continue near-real-time monitoring of the burden and VE of COVID-19 hospitalizations was lacking. Regular registration of all hospitalizations in the Netherlands, including International Classification of Diseases (ICD)-10 diagnosis codes, is provided by Dutch Hospital Data (DHD). These data are made available for data linkage and analyses within the Statistics Netherlands (CBS) remote access research environment some months after the end of the calendar year.

During the first quarter of 2023, circulation of SARS-CoV-2 increased, reaching a peak in early March [3]. In the third quarter of 2023, circulation rose gradually, followed by a more pronounced increase in the fourth quarter, driven by the emergence of the Omicron JN.1 variant. After a relatively high peak in the second half of December, SARS-CoV-2 circulation declined and remained at low levels throughout the spring of 2024. Circulation began to increase again in June, resulting in a summer wave that peaked in the second half of July. A second increase was observed in September which peaked early October 2024. Following this autumn wave, circulation decreased and remained at low levels during the winter of 2024.

We here describe the incidence of hospitalizations due to COVID-19 in 2023 and 2024 by age and medical risk group, and present vaccination coverage and VE of the 2023 and 2024 autumn vaccination rounds against COVID-19 hospitalization for persons aged 60 years and older in the Netherlands, by medical risk category.

## Methods

### Data sources

Expedited data on COVID-19 hospitalizations were provided by Dutch Hospital Data to the Statistics Netherlands remote access environment, which allows linkage to population registry data. In our primary analyses, only hospitalizations with an ICD-10 diagnosis code U07.1 or U07.2 (confirmed or suspected COVID-19) as primary or main diagnosis were defined as COVID-19 hospitalizations. VE analyses were repeated including all hospitalizations where U07.1 or U07.2 was included in the list of diagnoses.

The population register, including information on age, sex, and safety region (n=25) at individual level, was used to select the study population. Household socio-economic status was available as a composite measure, combining wealth, recent employment history and education level.

Medical risk groups were identified using healthcare utilization data from 2019 to 2023. The method for defining these groups is described in detail elsewhere [4]. In brief, individuals were classified as high, intermediate, or low risk for severe COVID-19 outcomes. Risk group classification was based on claims data for specialist healthcare consultations (sourced from Vektis) and prescription medication data at the ATC-4 code level (Anatomical Therapeutic Chemical classification). Diagnosis treatment combinations (DTCs) matching the conditions listed as high or intermediate risk were selected from the Vektis data, and, for certain conditions, this was supplemented with medication use data. The high-risk group included individuals who, according to available registration data, had received care or medication for conditions associated with a high risk of severe COVID-19 (e.g. hematologic malignancies, organ transplants) and were prioritized for the primary COVID-19 vaccination series. The intermediate-risk group included individuals with medical conditions that make them eligible for the seasonal influenza vaccination (e.g. persons with diabetes, cardiac disease), while all others were categorized as low medical risk. Infants below the age of 1 year were all assigned to the ‘low medical risk’ category as any possible chronic conditions were not likely captured by the 2019-2023 healthcare use data yet. Persons were allocated to a medical risk category based on their highest classification (i.e. if somebody had a condition in both the ‘high-risk’ group and the ‘intermediate risk’ group, they were assigned to the ‘high-risk’ group). Medical risk was assumed to be static (non-time-varying) during the study period.

### Study population

Vaccination coverage for both autumn vaccination rounds was determined using population denominators on 31 December of the corresponding year. For incidence estimates per age and medical risk group over 2022-2024, the population denominators were determined on 1 September 2023.

The VE analyses were restricted to the population aged 60 years or older, because this entire age-cohort was eligible for vaccination. For the VE analysis of the 2023 autumn vaccination round, the study population consisted of every person in the population registry aged 60 years or older on 25 September 2023, the start of the vaccination round. The end of the study period was 16 September 2024, when the 2024 autumn vaccination round started. For the VE 2024 analysis, the study period was 17 September 2024 to 31 December 2024, because 2025 hospitalization data were not yet available.

For VE estimates, only persons who had, since 2021, received at least one vaccination registered in the COVID-19 vaccination information and monitoring system (CIMS) were included, to reduce the risk of bias caused by non-consent for vaccination registration [5]. Persons with a COVID-19 hospitalization in the 90 days prior to the study period were excluded, as were persons who had received more booster doses than would be possible according to the vaccination program.

### Statistical analysis

Vaccination status was analyzed as a time-varying exposure, switching from ‘unvaccinated’ to ‘first 7 days’ on the date of vaccination, then to ‘vaccinated’ on day 8 post-vaccination. In the 2023 VE analysis, vaccinated person-time was divided into time since vaccination categories of 3 months up to 9+ months since vaccination. Persons were followed up until their first COVID-19 hospitalization, death, a subsequent COVID-19 vaccination during the study period, or the end of the study period, whichever came first.

Cox proportional hazards models were fitted, with calendar time as the underlying time scale. Age (in 5-year categories), sex, region (25 in total) and household socio-economic status (in quintiles) were added as covariates. VE was calculated as (1-Hazard Ratio)*100%. The assumed number needed to vaccinate (NNV) to prevent one hospitalization per year-quarter was estimated by first estimating the number of hospitalizations assumed to be avoided by vaccination, as the difference in expected and observed hospitalizations: N_avoided_ = VE * (N_hospitalizations_among_vaccinated_ * (1/(1-VE)). NNV was then calculated as NNV= 1/(N_avoided_ / person-year-quarters). Only the first year-quarter after the start of the autumn vaccination round was analyzed, to be able to compare 2023 and 2024 NNV estimates.

Analyses were performed in R version 4.4.3, using packages tidyverse, Epi, and survival. Results are based on calculations by the National Institute for Public Health and the Environment (RIVM) in project number 9248 using non-public microdata from Statistics Netherlands. Per the Statistics Netherlands policy, no numbers <10 or estimates based on <10 observations are presented.

## Results

### Incidence

In total, over 15,800 hospitalizations with COVID-19 as main or primary diagnosis were registered in 2023, and over 9.100 in 2024. The incidence of COVID-19 hospitalization over time in 2023-2024 peaked in the first and fourth quarter of 2023, and was lower in 2024 (Figure 1). Incidence was generally substantially higher in persons with a high medical risk, but the difference between medical risk groups was smaller in the age group of 80+ years. In absolute numbers (Figure S1), the intermediate medical risk group constituted the largest number of hospitalized patients (56% of hospitalizations). Persons with a low medical risk were not often hospitalized for COVID-19 in 2023 and 2024 (12% of hospitalizations). Infants under the age of 1 had a relatively large incidence of hospitalizations for COVID-19. The median duration of hospitalization of these infants was 1 day (IQR 1-2) during each year-quarter (Supplementary Table S5), compared to 3-5 days for age groups over 60 years old.

**Figure 1.**
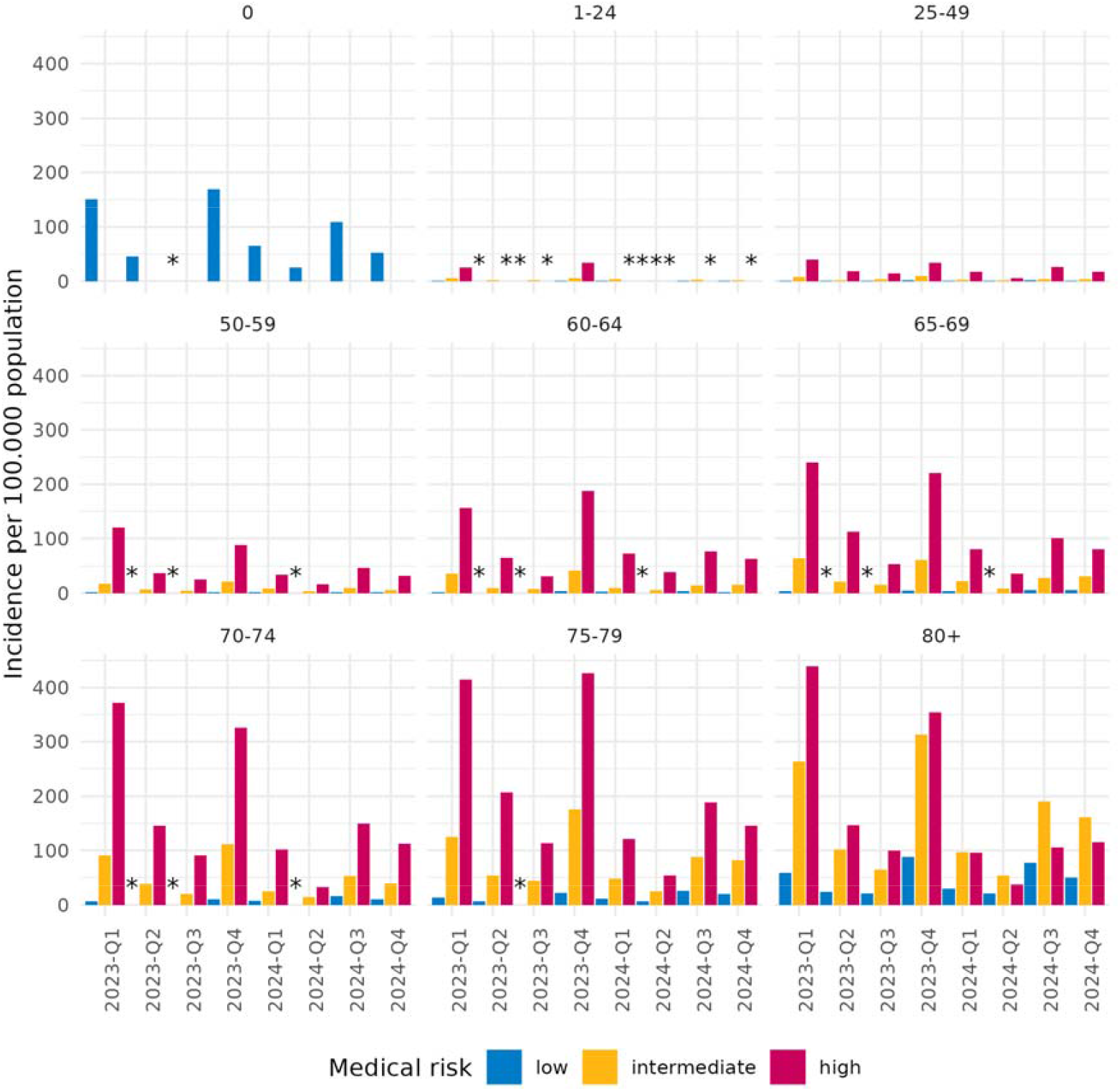
Incidence of hospitalizations with COVID-19 as main or primary diagnosis, by age group and medical risk group, 2023-2024. Infants below the age of 1 year were not divided into medical risk categories. * : Where no incidence is shown, this could be any incidence based on <10 events.. A version of this graph with a varying y axis is available as Supplementary Figure S3.

### Vaccination coverage

In both 2023 and 2024, COVID-19 vaccination was recommended for all persons aged 60 years and older, for people in intermediate and high medical risk groups and for people working in healthcare. Figure S2 shows vaccination coverage by age and medical risk group, measured on 31 December of each year, when the vaccination rounds had ended. Of all persons aged 60 years and older, 50.7% had a registered vaccination during the 2023 autumn vaccination round, and 47.2% during the 2024 autumn vaccination round. Vaccination coverage estimates did not differ substantially between low, intermediate and high medical risk groups above the age of 60 years.

### VE 2023 autumn round

VE of the 2023 autumn vaccination round was estimated at 67.9% (95%CI 59.0-74.8) in the first 3 months post-vaccination for persons aged 60 years or older with a low medical risk (Table 1). In the following year-quarters post-vaccination, this gradually decreased to 19.8% (95%CI -12.5-42.8%) 9-12 months post-vaccination. Similar VE was estimated for persons with an intermediate medical risk in this age group, but the VE for the highest medical risk group was initially lower (54.6% in the first 3 months post-vaccination, 95%CI 49.5-59.1%) and no remaining effectiveness was observed after 6 months post-vaccination. The estimated NNV was lowest for the high-risk group: only 28 persons aged 60 years and older needed to be vaccinated to prevent one hospitalization in the first 3 months post-vaccination. This NNV was estimated at 43 for the intermediate, and 276 for the low risk group. Over the entire study period of the 2023 vaccination campaign (25 September 2023 to 16 September 2024) around 3,300 hospitalizations are estimated to have been prevented by this vaccination round. The sensitivity analysis including all hospitalizations with COVID-19 listed among the diagnosis codes showed similar trends, with only moderately lowered VE estimates (Supplementary Table S3). A substantially lower risk of hospitalization was also observed in the first 7 days post-vaccination.

**Table 1.** Hospitalizations with COVID-19 as main or primary diagnosis, person-years included in the analysis, incidence, estimated vaccine effectiveness with 95% confidence interval, estimated number of avoided hospitalizations and number needed to vaccinate to prevent one hospitalization, 2023 autumn vaccination round, among persons aged 60 years and older by medical risk group, 25 September 2023-16 September 2024.

| Medical risk | Vaccination status autumn 2023 | Person-years | COVID-19 hospitalizations | VE (95%CI) | Avoided COVID-19 hospitalizations | NNV |
| --- | --- | --- | --- | --- | --- | --- |
| low | not vaccinated | 799983 | 415 | [ref] |  |  |
| low | first 7 days post-vaccination | 16544 | <10 |  |  |  |
| low | vaccinated (7d-3m) | 219084 | 94 | 67.9% (59.0-74.8%) | 199 | 275 |
| low | vaccinated (3-6m) | 218573 | 24 | 59.3% (34.7-74.7%) | 35 | 1563 |
| low | vaccinated (6-9m) | 217450 | 98 | 29.1% (6.5-46.2%) | 40 | 1352 |
| low | vaccinated (9-12m) | 86002 | 64 | 19.9% (-12.4-42.9%) | 16 | 1352 |
| intermediate | not vaccinated | 920015 | 3550 | [ref] |  |  |
| intermediate | first 7 days post-vaccination | 23022 | 104 | 50.2% (39.3-59.1%) | 105 | 55 |
| intermediate | vaccinated (7d-3m) | 302463 | 935 | 65.3% (62.5-67.9%) | 1760 | 43 |
| intermediate | vaccinated (3-6m) | 299499 | 143 | 55.9% (46.5-63.7%) | 181 | 413 |
| intermediate | vaccinated (6-9m) | 296638 | 625 | 24.7% (15.9-32.7%) | 205 | 362 |
| intermediate | vaccinated (9-12m) | 121198 | 332 | 21.0% (8.6-31.7%) | 88 | 343 |
| high | not vaccinated | 217884 | 1894 | [ref] |  |  |
| high | first 7 days post-vaccination | 5388 | 48 | 54.9% (39.8-66.2%) | 58 | 23 |
| high | vaccinated (7d-3m) | 69419 | 510 | 54.6% (49.6-59.2%) | 613 | 28 |
| high | vaccinated (3-6m) | 65809 | 88 | 27.4% (6.3-43.8%) | 33 | 495 |
| high | vaccinated (6-9m) | 63572 | 245 | 1.1% (-18.5-17.4%) | 3 | 5832 |
| high | vaccinated (9-12m) | 26088 | 119 | -9.7% (-40.0-14.1%) | -11 | -620 |

### VE 2024 autumn round

The short-term (up to 3 months post-vaccination) VE of the 2024 autumn vaccination round up to 31 December 2024 was estimated at 61.7% for persons aged 60 years or older with low medical risk (95%CI 44.4-73.7%) and was again similar for the intermediate risk group (61.8%, 95%CI 55.5-67.2%, Table 2). The high-risk group aged 60 years and older had an estimated VE of 54.9% (95%CI 42.6-64.5%). The estimated NNV in these first months after the start of the campaign was 93 for the high-risk group, 138 and 625 for the intermediate and low risk groups, respectively. Between 17 September and 31 December 2024, around 500 hospitalizations were estimated to have been prevented by vaccination. The sensitivity analysis including all hospitalizations with a COVID-19 diagnosis code resulted in VE estimates of 5-8 percentage points lower only for low and intermediate medical risk groups (Supplementary Table S4).

**Table 2.**
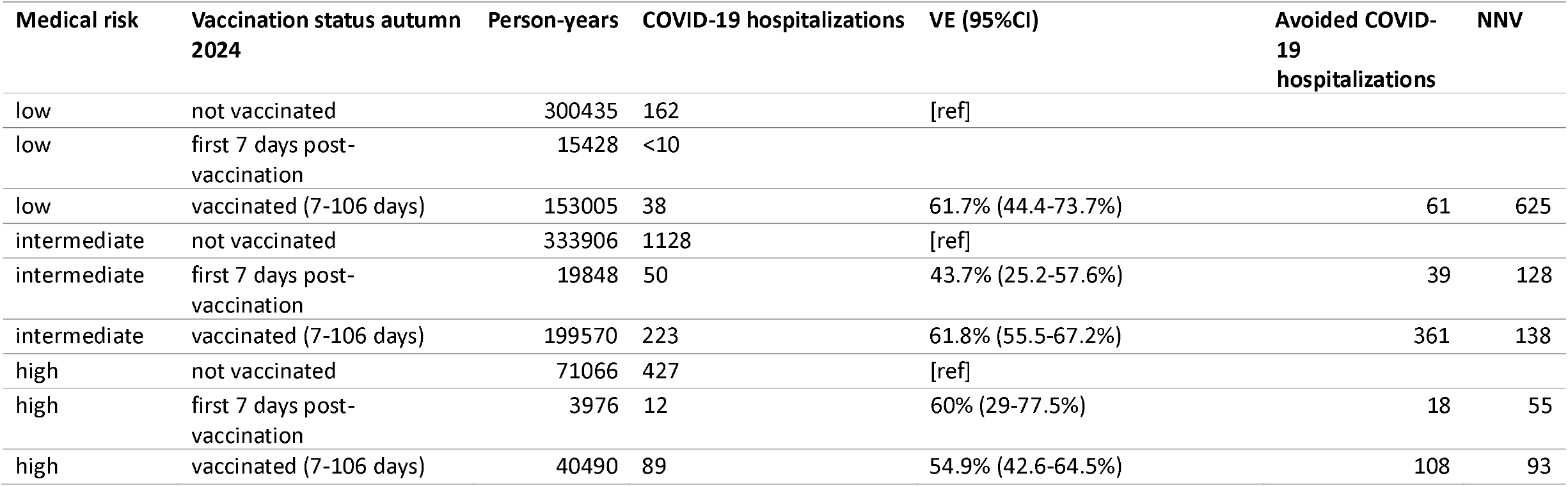
Hospitalizations with COVID-19 as main or primary diagnosis, person-years included in the analysis, incidence, estimated vaccine effectiveness with 95% confidence interval, estimated number of avoided hospitalizations and number needed to vaccinate to prevent one hospitalization, 2024 autumn vaccination round, among persons aged 60 years and older, 17 September – 31 December 2024.

| Medical risk | Vaccination status autumn 2024 | Person-years | COVID-19 hospitalizations | VE (95%CI) | Avoided COVID-19 hospitalizations | NNV |
| --- | --- | --- | --- | --- | --- | --- |
| low | not vaccinated | 300435 | 162 | [ref] |  |  |
| low | first 7 days post-vaccination | 15428 | <10 |  |  |  |
| low | vaccinated (7-106 days) | 153005 | 38 | 61.7% (44.4-73.7%) | 61 | 625 |
| intermediate | not vaccinated | 333906 | 1128 | [ref] |  |  |
| intermediate | first 7 days post-vaccination | 19848 | 50 | 43.7% (25.2-57.6%) | 39 | 128 |
| intermediate | vaccinated (7-106 days) | 199570 | 223 | 61.8% (55.5-67.2%) | 361 | 138 |
| high | not vaccinated | 71066 | 427 | [ref] |  |  |
| high | first 7 days post-vaccination | 3976 | 12 | 60% (29-77.5%) | 18 | 55 |
| high | vaccinated (7-106 days) | 40490 | 89 | 54.9% (42.6-64.5%) | 108 | 93 |

## Discussion

In 2023-2024, over 24.000 hospitalizations with COVID-19 as main or primary diagnosis were registered in the Netherlands. The incidence of COVID-19 hospitalization was highest among high and intermediate medical risk groups aged 60 years and older. These incidences occurred despite vaccination campaigns targeting these groups in the autumns of 2022, 2023 and 2024. A substantial burden was preventable, as autumn boosters continued to offer protection against COVID-19 hospitalization and vaccination coverage was suboptimal. Infants below the age of 1 year had a high incidence of COVID-19 hospitalizations. However, the median duration of hospitalization of these infants was 1 day, suggesting that these might be largely precautionary hospitalizations of infants with a SARS-CoV-2 infection rather than cases of severe COVID-19.

The estimates of the VE against hospitalisation over time since vaccination for the 2023 autumn booster presented in this study show the VE is not long-lasting. Previously, we estimated VE of the 2023 autumn booster between 9 October and 5 December 2023 at 70.7%, based on the NICE data with a limited number of contributing hospitals. Our current estimates, based on more complete data, are a bit lower for the 0-3 months post-vaccination period. The difference in these estimates could be due to a longer time since vaccination in the current study, especially for the medical high-risk group. Among this high-risk group, the decrease in VE indicated no remaining protection after 6 months post-vaccination. The decreasing effectiveness over time, combined with the unpredictable seasonality of COVID-19 incidence waves, complicates determining the optimal timing of COVID-19 vaccination rounds. This may indicate benefit of a spring booster round for the highest risk groups, as is recommended in the United Kingdom [6]. The lower estimated NNV illustrates that despite a lower VE, the high-risk group benefits most per vaccinated individual. For the low and intermediate risk groups, our results suggest a low to moderate degree of protection remains up to 9-12 months post-vaccination.

For the 2024 autumn vaccination round, short-term VE was moderate, at around 53-56%. A recently published estimate of the autumn 2024 JN.1 VE against COVID-19 hospitalization, in October 2024-January 2025 among six European study sites, was comparable, between 53.7 and 60.3% among persons aged 60 years and older, with a time since vaccination between 14 and 119 days [7]. A population-wide cohort study from Denmark in the same study period found a higher VE of 70-85% for the two vaccines used [8]. Test-negative case-control studies, in England and the United States, found a lower VE against COVID-19 hospitalization in persons aged 65 years and older (around 45% in the first 3 months since vaccination) [9-11]. A test-negative case-control study from the United States with longer follow-up time found a lower and no longer significant VE against COVID-19 hospitalization of the 2024 autumn round after > 180 days post-vaccination [11].

Already in the first 7 days after vaccine administration, a much lower risk of COVID-19 hospitalization was observed compared to unvaccinated person-time. Several aspects may contribute to this phenomenon. Firstly, we took the date of hospitalization as event date, while infection, and likely disease onset, occurred earlier and patients likely did not get vaccinated in the days between disease onset and hospitalization. Also, the booster response after several previous exposures (vaccine doses and infections) may occur more rapidly than the commonly assumed 7 days [12]. Further, the lower risk of hospitalization in a time period where the vaccine is not expected to have a large effect yet, may indicate a substantial healthy vaccinee bias, where persons in relatively good health are more likely to choose vaccination. This unmeasured confounding could also cause upward bias in VE estimates after >7 days post-vaccination [4, 13].

Our study has several limitations. Importantly, the classification of medical risk groups is flawed. We based this classification on available data on specialized medical healthcare use and medication data for 2019-2023, but not all relevant chronic conditions can be captured by this method [4]. The extent of this misclassification is unknown. In 2021, persons with an immunocompromising condition were eligible for a third dose in the primary series. Of all persons who received such a third primary dose according to CIMS, only 60% was classified as belonging to the medical high-risk group by our method. Likely, this misclassification diluted the difference between the risk groups and the difference in incidences may be larger in reality.

The requirement of informed consent for registration of COVID-19 vaccinations is another important limitation of our study. The resulting misclassification of vaccinees as unvaccinated during the campaign possibly led to an underestimation of VE, despite 98% of administered vaccinations being registered in the 2023 and 2024 autumn campaigns [5]. We attempted to reduce this bias by including only persons who had at least one vaccination registered in CIMS and, therefore, were more likely to consent to registration of (further) vaccinations.

Because we did not have data on SARS-CoV-2 infections that did not lead to hospitalization, we could not account for infection-induced immunity in our analysis. Therefore, some of the observed waning effectiveness may be spurious, caused by a differential reduction of susceptibility if recent infection-induced immunity was accrued more quickly by persons not vaccinated during the 2023 autumn campaign [14]. Also, changes in circulating SARS-CoV-2 variants may have affected VE over time, separate from waning effectiveness, although studies from Denmark and the United States did not observe clear differences in VE against hospitalization for variants circulating in 2024 [8, 11].

In conclusion, despite increasing population immunity and yearly vaccination rounds, still a significant disease burden of COVID-19 hospitalizations exists in the Netherlands. Part of this disease burden could be averted by increasing vaccine uptake. As VE decreases over time since vaccination, revaccination likely remains beneficial, mostly for intermediate and high medical risk groups.

## Data Availability

Results based on calculations by the RIVM in project number 9248 using non-public microdata from Statistics Netherlands. Under certain conditions, these microdata are accessible for statistical and scientific research. For further information: https://www.cbs.nl/en-gb/our-services/customised-services-microdata/microdata-conducting-your-own-research

