## Supplementary material for "COVID-19 hospitalizations in the Netherlands, 2023-2024: disease burden and vaccine effectiveness": Suppl

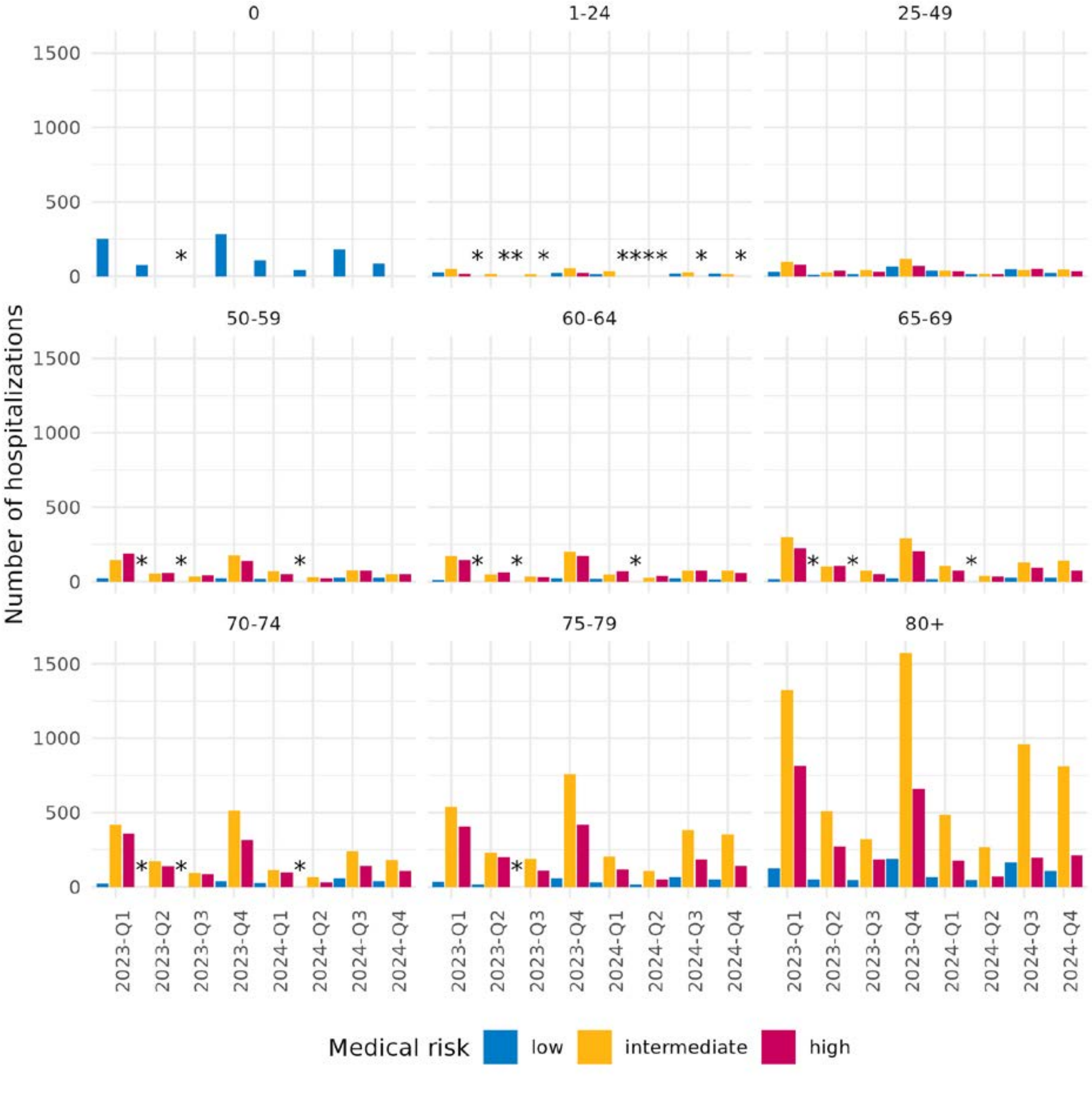


**Figure S1.** Number of hospitalizations with COVID-19 as main or primary diagnosis, by age group and medical risk group, 2023-2024. *: Where no hospitalizations are shown, this can be any number <10.

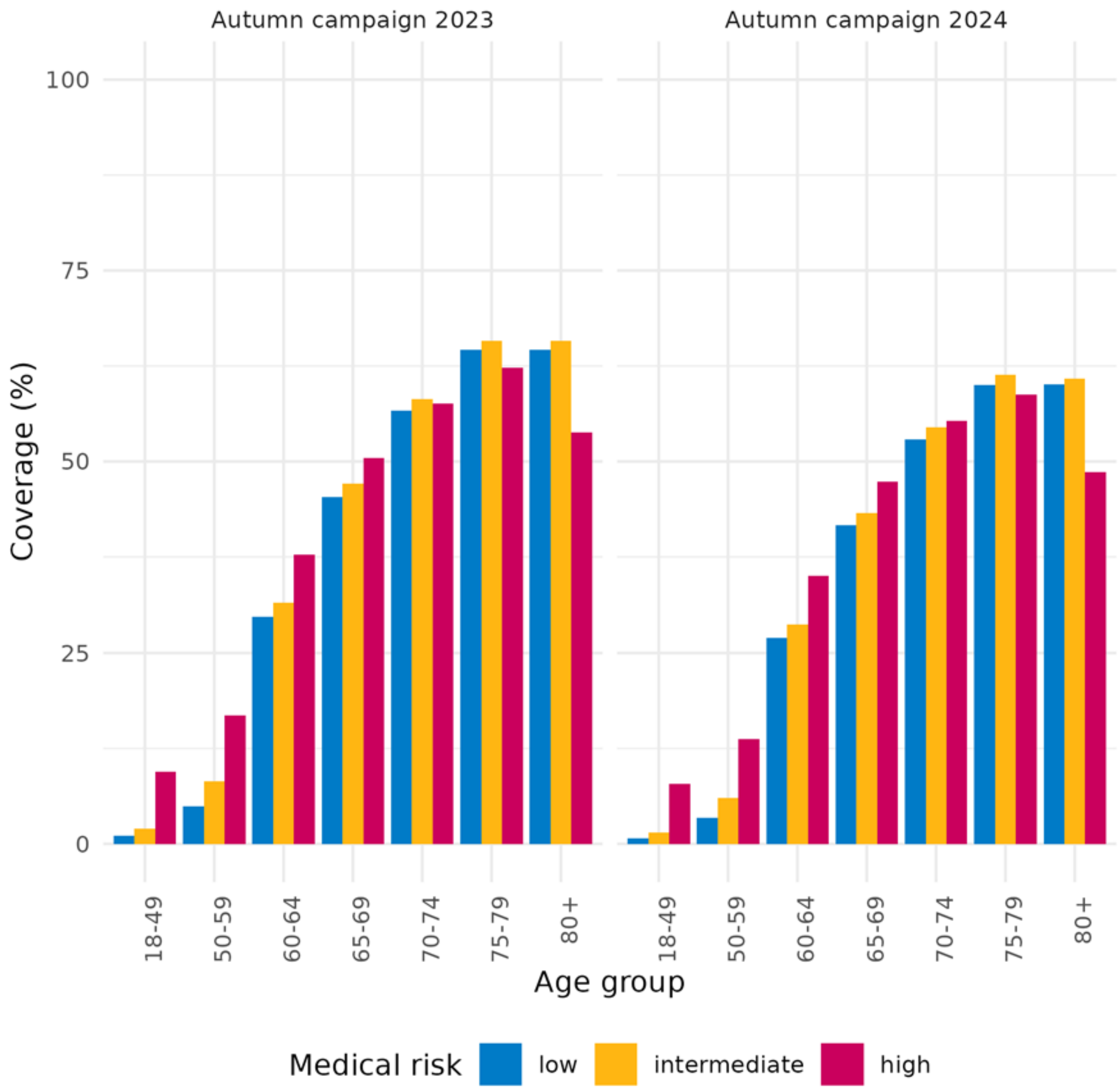


**Figure S2.** COVID-19 vaccination coverage, measured on December 31, 2023 and 2024, by age and medical risk group.

**Table S1.** Study population characteristics for VE estimates of the 2023 autumn vaccination round against hospitalization with COVID-19 as main or primary diagnosis.

| | | **Unvaccinated** | **Vaccinated** |
|---|---|---|---|
| **n** | | 1897164 | 2396809 |
| **Age group (years)** | 60-64 | 575150 (30.3) | 321383 (13.4) |
| | 65-69 | 469305 (24.7) | 480276 (20.0) |
| | 70-74 | 329560 (17.4) | 525251 (21.9) |
| | 75-79 | 237146 (12.5) | 516239 (21.5) |
| | 80-84 | 141975 ( 7.5) | 311046 (13.0) |
| | 85-89 | 89045 ( 4.7) | 169943 ( 7.1) |
| | 90+ | 54983 ( 2.9) | 72671 ( 3.0) |
| **Sex** | Male | 891035 (47.0) | 1152569 (48.1) |
| | Female | 1006129 (53.0) | 1244240 (51.9) |

| | | | |
|---|---|---|---|
| **Medical risk group** | Low | 775499 (40.9) | 889076 (37.1) |
| | Intermediate | 891219 (47.0) | 1227070 (51.2) |
| | High | 230446 (12.1) | 280663 (11.7) |
| **Household socio-economic status (quintiles)** | 1 | 397265 (20.9) | 311837 (13.0) |
| | 2 | 479958 (25.3) | 690196 (28.8) |
| | 3 | 422297 (22.3) | 685027 (28.6) |
| | 4 | 325100 (17.1) | 451737 (18.8) |
| | 5 | 194271 (10.2) | 192243 ( 8.0) |
| | NA | 78273 ( 4.1) | 65769 ( 2.7) |
| **Region** | Amsterdam-Amstelland | 93750 ( 5.0) | 96349 ( 4.0) |
| | Brabant-Noord | 74766 ( 3.9) | 103312 ( 4.3) |
| | Brabant-Zuidoost | 83698 ( 4.4) | 117925 ( 4.9) |
| | Drenthe | 60374 ( 3.2) | 76331 ( 3.2) |
| | Flevoland | 39219 ( 2.1) | 44231 ( 1.8) |
| | Fryslân | 89064 ( 4.7) | 84961 ( 3.6) |
| | Gelderland-Midden | 69863 ( 3.7) | 100779 ( 4.2) |
| | Gelderland-Zuid | 54807 ( 2.9) | 83215 ( 3.5) |
| | Gooi en Vechtstreek | 25373 ( 1.3) | 37126 ( 1.6) |
| | Groningen | 62166 ( 3.3) | 84122 ( 3.5) |
| | Haaglanden | 105625 ( 5.6) | 132424 ( 5.5) |
| | Hollands-Midden | 82663 ( 4.4) | 119152 ( 5.0) |
| | IJsselland | 57452 ( 3.0) | 70440 ( 2.9) |
| | Kennemerland | 56576 ( 3.0) | 76033 ( 3.2) |
| | Limburg-Noord | 71702 ( 3.8) | 83505 ( 3.5) |
| | Limburg-Zuid | 85971 ( 4.5) | 93307 ( 3.9) |
| | Midden- en West-Brabant | 120506 ( 6.4) | 174716 ( 7.3) |
| | Noord- en Oost-Gelderland | 100693 ( 5.3) | 135582 ( 5.7) |
| | Noord-Holland-Noord | 83299 ( 4.4) | 97944 ( 4.1) |
| | Rotterdam-Rijnmond | 136919 ( 7.2) | 151635 ( 6.3) |
| | Twente | 81454 ( 4.3) | 80088 ( 3.3) |
| | Utrecht | 125802 ( 6.6) | 173130 ( 7.2) |
| | Zaanstreek-Waterland | 40762 ( 2.2) | 43533 ( 1.8) |
| | Zeeland | 43023 ( 2.3) | 66171 ( 2.8) |
| | Zuid-Holland-Zuid | 48150 ( 2.5) | 65140 ( 2.7) |

**Table S2.** Study population characteristics for VE estimates of the 2024 autumn vaccination round against hospitalization with COVID-19 as main or primary diagnosis.

| | | **Unvaccinated** | **Vaccinated** |
|---|---|---|---|
| **n** | | 2216905 | 2320181 |
| **Age group (years)** | 60-64 | 726701 (32.8) | 341170 (14.7) |
| | 65-69 | 514410 (23.2) | 453783 (19.6) |
| | 70-74 | 358055 (16.2) | 493447 (21.3) |

| | | | |
|---|---|---|---|
| | 75-79 | 279749 (12.6) | 497229 (21.4) |
| | 80-84 | 169441 ( 7.6) | 304156 (13.1) |
| | 85-89 | 105919 ( 4.8) | 162980 ( 7.0) |
| | 90+ | 62630 ( 2.8) | 67416 ( 2.9) |
| **Sex** | Male | 1049103 (47.3) | 1118275 (48.2) |
| | Female | 1167802 (52.7) | 1201906 (51.8) |
| **Medical risk group** | Low | 956475 (43.1) | 907799 (39.1) |
| | Intermediate | 1031139 (46.5) | 1175105 (50.6) |
| | High | 229291 (10.3) | 237277 (10.2) |
| **Household socio-economic status (quintiles)** | 1 | 434220 (19.6) | 279530 (12.0) |
| | 2 | 528898 (23.9) | 633190 (27.3) |
| | 3 | 487510 (22.0) | 658808 (28.4) |
| | 4 | 417491 (18.8) | 466066 (20.1) |
| | 5 | 269847 (12.2) | 235184 (10.1) |
| | NA | 78939 ( 3.6) | 47403 ( 2.0) |
| **Region** | Amsterdam-Amstelland | 107950 ( 4.9) | 94189 ( 4.1) |
| | Brabant-Noord | 86974 ( 3.9) | 101971 ( 4.4) |
| | Brabant-Zuidoost | 98882 ( 4.5) | 114021 ( 4.9) |
| | Drenthe | 75176 ( 3.4) | 77852 ( 3.4) |
| | Flevoland | 46799 ( 2.1) | 42918 ( 1.8) |
| | Fryslân | 101551 ( 4.6) | 81482 ( 3.5) |
| | Gelderland-Midden | 81769 ( 3.7) | 98847 ( 4.3) |
| | Gelderland-Zuid | 65814 ( 3.0) | 80984 ( 3.5) |
| | Gooi en Vechtstreek | 29913 ( 1.3) | 35785 ( 1.5) |
| | Groningen | 74134 ( 3.3) | 79321 ( 3.4) |
| | Haaglanden | 125159 ( 5.6) | 126659 ( 5.5) |
| | Hollands-Midden | 99441 ( 4.5) | 114872 ( 5.0) |
| | IJsselland | 65237 ( 2.9) | 70390 ( 3.0) |
| | Kennemerland | 67620 ( 3.1) | 72645 ( 3.1) |
| | Limburg-Noord | 83659 ( 3.8) | 80051 ( 3.5) |
| | Limburg-Zuid | 100579 ( 4.5) | 87102 ( 3.8) |
| | Midden- en West-Brabant | 145152 ( 6.5) | 166558 ( 7.2) |
| | Noord- en Oost-Gelderland | 115592 ( 5.2) | 134226 ( 5.8) |
| | Noord-Holland-Noord | 95389 ( 4.3) | 95514 ( 4.1) |
| | Rotterdam-Rijnmond | 159886 ( 7.2) | 144021 ( 6.2) |
| | Twente | 89402 ( 4.0) | 80558 ( 3.5) |
| | Utrecht | 144485 ( 6.5) | 173283 ( 7.5) |
| | Zaanstreek-Waterland | 46675 ( 2.1) | 42510 ( 1.8) |
| | Zeeland | 53320 ( 2.4) | 61182 ( 2.6) |
| | Zuid-Holland-Zuid | 56347 ( 2.5) | 63240 ( 2.7) |

**Table S3.** Hospitalizations with COVID-19 as any diagnosis, person-years included in the analysis, incidence, vaccine effectiveness with 95% confidence interval, number of avoided hospitalizations and number needed to vaccinate to prevent one hospitalization, 2023 autumn vaccination round, among persons aged 60 years and older, September 2023- September 2024.

| Medical risk | Vaccination status autumn 2023 | Person-years | COVID-19 hospitalizations | VE (95%CI) | Avoided COVID-19 hospitalizations | NNV |
|---|---|---|---|---|---|---|
| low | not vaccinated | 799859 | 723 | [ref] | | |
| low | first 7 days post-vaccination | 16544 | 12 | 54.7% (19.3-74.6%) | 14 | 285 |
| low | vaccinated (7d-3m) | 219076 | 177 | 62.6% (55.1-68.8%) | 296 | 185 |
| low | vaccinated (3-6m) | 218553 | 46 | 60.5% (44.5-71.9%) | 70 | 775 |
| low | vaccinated (6-9m) | 217423 | 165 | 33.1% (17.6-45.6%) | 82 | 666 |
| low | vaccinated (9+m) | 85984 | 113 | 21.8% (-0.5-39.1%) | 32 | 682 |
| intermediate | not vaccinated | 918847 | 5968 | [ref] | | |
| intermediate | first 7 days post-vaccination | 23020 | 169 | 51.1% (43-58.1%) | 177 | 33 |
| intermediate | vaccinated (7d-3m) | 302353 | 1699 | 61.8% (59.5-64%) | 2749 | 28 |
| intermediate | vaccinated (3-6m) | 299310 | 314 | 48.7% (41.4-55.2%) | 298 | 251 |
| intermediate | vaccinated (6-9m) | 296413 | 1013 | 21.8% (14.6-28.4%) | 282 | 262 |
| intermediate | vaccinated (9+m) | 121067 | 558 | 18.2% (8.4-26.9%) | 124 | 244 |
| high | not vaccinated | 217383 | 3115 | [ref] | | |
| high | first 7 days post-vaccination | 5388 | 92 | 46.5% (34.1-56.6%) | 80 | 17 |
| high | vaccinated (7d-3m) | 69369 | 846 | 53.8% (49.9-57.5%) | 985 | 18 |
| high | vaccinated (3-6m) | 65737 | 167 | 23.9% (8.3-36.9%) | 52 | 313 |
| high | vaccinated (6-9m) | 63486 | 388 | -0.1% (-15.6-13.3%) | 0 | -40947 |
| high | vaccinated (9+m) | 26043 | 191 | -9.8% (-33.2-9.5%) | -17 | -382 |

**Table S4.** Hospitalizations with COVID-19 as any diagnosis, person-years included in the analysis, incidence, vaccine effectiveness with 95% confidence interval, number of avoided hospitalizations and number needed to vaccinate to prevent one hospitalization, 2024 autumn vaccination round, among persons aged 60 years and older, September - December 2024.

| Medical risk | Vaccination status autumn 2024 | Person-years | COVID-19 hospitalizations | VE (95%CI) | Avoided COVID-19 hospitalizations | NNV |
|---|---|---|---|---|---|---|
| **low** | not vaccinated | 300405 | 352 | [ref] | | |
| low | first 7 days post-vaccination | 15428 | <10 | | | |
| low | vaccinated (7+ days) | 152998 | 87 | 53.5% (40.3-63.7%) | 100 | 382 |
| intermediate | not vaccinated | 333797 | 1884 | [ref] | | |
| intermediate | first 7 days post-vaccination | 19847 | 82 | 44.1% (30.2-55.2%) | 65 | 77 |
| intermediate | vaccinated (7+ days) | 199540 | 426 | 56.4% (51.2-61%) | 551 | 91 |
| high | not vaccinated | 71030 | 685 | [ref] | | |
| high | first 7 days post-vaccination | 3975 | 23 | 52.1% (27.3-68.4%) | 25 | 40 |
| high | vaccinated (7+ days) | 40481 | 140 | 55.7% (46.4-63.5%) | 176 | 57 |

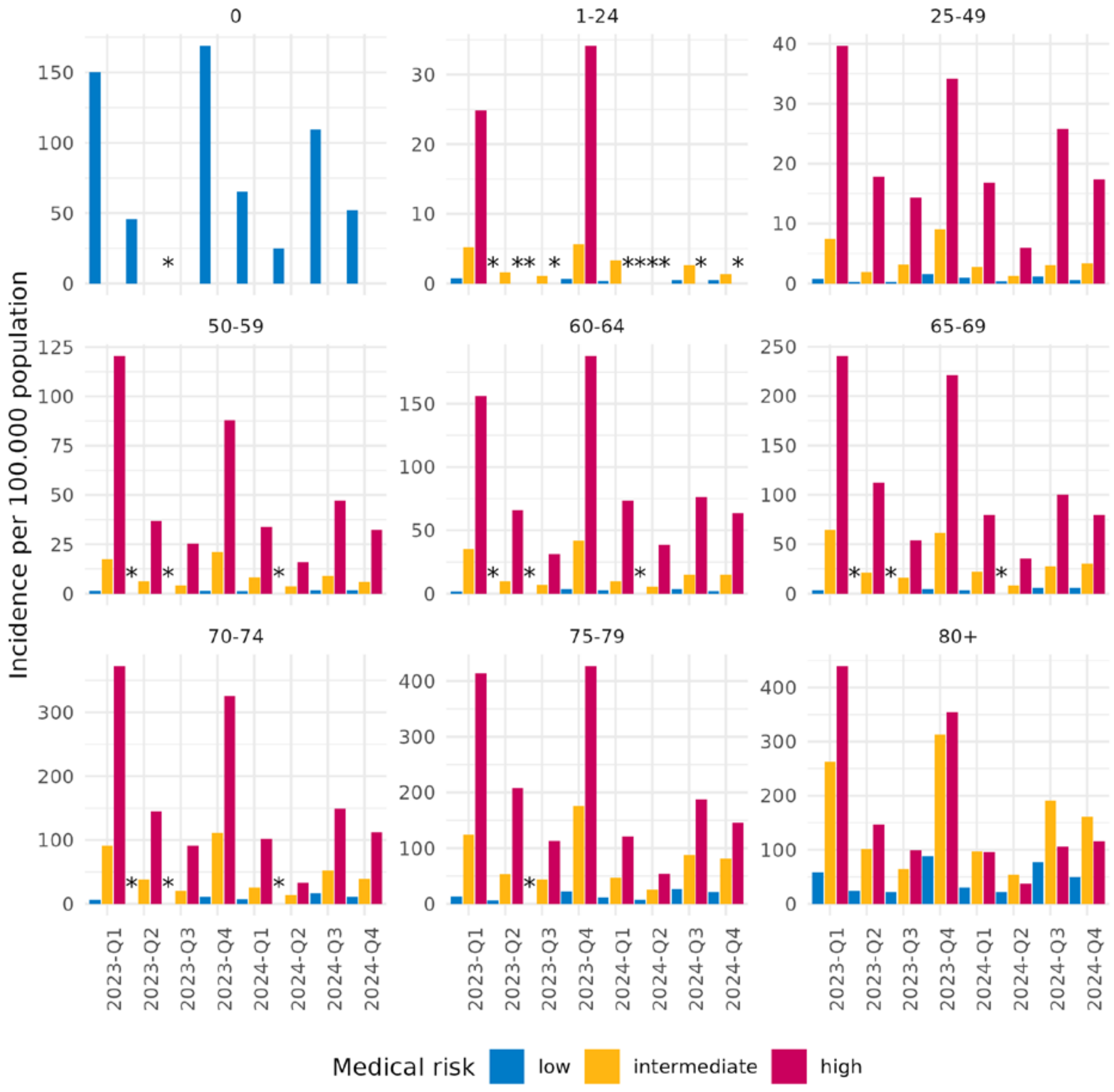


**Figure S3.** Incidence of hospitalizations with COVID-19 as main or primary diagnosis, by age group and medical risk group, 2023-2024. * : Where no incidence is shown, this could be any incidence based on <10 events. Infants below the age of 1 year were not divided into medical risk categories.

**Table S5.** Duration of hospitalizations with COVID-19 as main or primary diagnosis, by age group and year-quarter, 2023-2024.

| Age group | Year-quarter | COVID-19 hospitalizations | Duration in days (median (IQR)) |
|---|---|---|---|
| 0 | 2023-Q1 | 264 | 1 (1-2) |
| 0 | 2023-Q2 | 88 | 1 (1-2) |
| 0 | 2023-Q3 | 77 | 1 (1-2) |
| 0 | 2023-Q4 | 291 | 1 (1-2) |
| 0 | 2024-Q1 | 114 | 1 (1-2) |
| 0 | 2024-Q2 | 44 | 1 (1-2) |

| | | | |
|---|---|---|---|
| 0 | 2024-Q3 | 182 | 1 (1-2) |
| 0 | 2024-Q4 | 87 | 1 (1-2) |
| 1-24 | 2023-Q1 | 94 | 2 (1-3) |
| 1-24 | 2023-Q2 | 25 | 1 (1-3) |
| 1-24 | 2023-Q3 | 26 | 1 (1-2) |
| 1-24 | 2023-Q4 | 100 | 2 (1-3) |
| 1-24 | 2024-Q1 | 53 | 3 (2-6) |
| 1-24 | 2024-Q2 | 17 | 2 (1-3) |
| 1-24 | 2024-Q3 | 49 | 2 (1-4) |
| 1-24 | 2024-Q4 | 39 | 2 (1-3) |
| 25-49 | 2023-Q1 | 209 | 2 (1-4) |
| 25-49 | 2023-Q2 | 74 | 2 (1-5) |
| 25-49 | 2023-Q3 | 82 | 2 (1-6) |
| 25-49 | 2023-Q4 | 258 | 2 (1-3) |
| 25-49 | 2024-Q1 | 108 | 2 (1-4) |
| 25-49 | 2024-Q2 | 43 | 2 (1-3) |
| 25-49 | 2024-Q3 | 139 | 2 (1-3) |
| 25-49 | 2024-Q4 | 101 | 2 (1-4) |
| 50-59 | 2023-Q1 | 359 | 3 (2-8) |
| 50-59 | 2023-Q2 | 119 | 4 (2-6) |
| 50-59 | 2023-Q3 | 80 | 2 (1-3) |
| 50-59 | 2023-Q4 | 338 | 3 (2-5) |
| 50-59 | 2024-Q1 | 143 | 3 (2-6) |
| 50-59 | 2024-Q2 | 64 | 3 (2-5) |
| 50-59 | 2024-Q3 | 177 | 3 (2-5) |
| 50-59 | 2024-Q4 | 128 | 3 (2-5) |
| 60-64 | 2023-Q1 | 331 | 4 (2-7) |
| 60-64 | 2023-Q2 | 119 | 3 (2-6) |
| 60-64 | 2023-Q3 | 69 | 3 (2-8) |
| 60-64 | 2023-Q4 | 402 | 3 (2-6) |
| 60-64 | 2024-Q1 | 135 | 4 (2-7) |
| 60-64 | 2024-Q2 | 69 | 4 (2-7) |
| 60-64 | 2024-Q3 | 166 | 3 (1-6) |
| 60-64 | 2024-Q4 | 145 | 3 (2-6) |
| 65-69 | 2023-Q1 | 548 | 4 (2-8) |
| 65-69 | 2023-Q2 | 210 | 4 (2-7) |
| 65-69 | 2023-Q3 | 136 | 3 (2-5) |
| 65-69 | 2023-Q4 | 521 | 3 (2-6) |
| 65-69 | 2024-Q1 | 196 | 4 (2-7) |
| 65-69 | 2024-Q2 | 77 | 3 (2-6) |
| 65-69 | 2024-Q3 | 249 | 3 (2-6) |
| 65-69 | 2024-Q4 | 244 | 4 (2-6) |
| 70-74 | 2023-Q1 | 817 | 4 (2-8) |
| 70-74 | 2023-Q2 | 330 | 4 (2-8) |
| 70-74 | 2023-Q3 | 196 | 4 (2-9) |
| 70-74 | 2023-Q4 | 879 | 4 (2-7) |
| 70-74 | 2024-Q1 | 242 | 4 (2-8) |
| 70-74 | 2024-Q2 | 107 | 3 (2-6) |
| 70-74 | 2024-Q3 | 445 | 3 (2-6) |
| 70-74 | 2024-Q4 | 327 | 4 (2-7) |
| 75-79 | 2023-Q1 | 995 | 4 (2-8) |
| 75-79 | 2023-Q2 | 459 | 4 (2-8) |

| | | | |
|---|---|---|---|
| 75-79 | 2023-Q3 | 313 | 4 (2-7) |
| 75-79 | 2023-Q4 | 1259 | 4 (2-7) |
| 75-79 | 2024-Q1 | 355 | 4 (2-8) |
| 75-79 | 2024-Q2 | 180 | 4 (2-7) |
| 75-79 | 2024-Q3 | 633 | 4 (2-7) |
| 75-79 | 2024-Q4 | 549 | 4 (2-7) |
| 80+ | 2023-Q1 | 2301 | 5 (3-9) |
| 80+ | 2023-Q2 | 861 | 5 (3-9) |
| 80+ | 2023-Q3 | 556 | 5 (2-9) |
| 80+ | 2023-Q4 | 2450 | 4 (2-8) |
| 80+ | 2024-Q1 | 730 | 5 (2-8) |
| 80+ | 2024-Q2 | 383 | 4 (2-8) |
| 80+ | 2024-Q3 | 1321 | 4 (3-8) |
| 80+ | 2024-Q4 | 1133 | 4 (2-8) |
